# Association of comorbidities and socioeconomic deprivation among people who died from dementia in England Between 2013-2023: analysis of death certificates

**DOI:** 10.1101/2025.06.24.25330177

**Authors:** Sedigheh Zabihi, Michael Jackson, Elizabeth L Sampson, Sube Banerjee, Charlotte Kenten, Claudia Cooper

## Abstract

**Objective:** To describe the sociodemographic characteristics and comorbidities of people who died from dementia between 2013 and 2023 in England.

**Methods:** We analysed death certificates reported in England from 2013-23. We report the number, age, sex, country of origin and socioeconomic status of people who died with a dementia diagnosis recorded (as a primary or contributory cause); the dementia subtype diagnoses and recorded comorbidities. We tested the hypothesis that number of comorbid disorders would be higher in more deprived areas.

**Results:** There were 987 719 certificates in this period that recorded dementia as a cause of death, of which 693 663 (70.2%) recorded dementia as the primary cause of death. 62.2% (n=614 419) of those who died from dementia were women, 65% (n=643 026) were aged 85 and over, and most (846 584, 85.7%) were born in England. Fifteen percent (n=149 447) of included death certificates recorded dementia as sole cause of death; others included up to ten other contributory conditions (median=1; IQR:1-2), of which influenza and pneumonia (183 203; 18.5%), ischaemic heart diseases (114 871; 11.6%), cancers (107 444; 10.8%), hypertensive diseases (99 517; 10%) and diabetes (98 517; 10%) were the most common. After controlling for age and sex, death certificates of people living in areas with higher deprivation included a higher number of comorbidities (R^2^=0.01, p<0.001)

**Conclusions:** Policies to reduce inequities in dementia care need to account for the more complex health needs of people with dementia living in more deprived areas towards the end of life.

## Introduction

Most people who die from dementia have co-existing physical and mental comorbidities, which are associated with more rapid cognitive and functional decline, and lower quality of life [1]. They place a significant burden on the nation’s health, healthcare use, and social care services [2]. The UK government’s 2025 health mission plans to tackle persistent inequalities in health by shortening the amount of time people spend in ill-health and earlier identification and management of chronic conditions [3]. Knowing more about the comorbidities people with dementia experience in their final years, and who is most at risk within the population could support delivery of this mission, by informing how policymakers plan and prioritise services. Death certificates are an important source of information about people who die with dementia, as they can avoid bias related to the lesser likelihood of more deprived populations taking part in research in this area [4].

The cause of death in a person with advanced dementia commonly involves pneumonia, because neurodegenerative processes diminish coughing and swallowing abilities, increasing the likelihood of aspiration and subsequent pneumonia. Such illnesses can be conceptualised as consequences of end-stage dementia. Some conditions, such as cardiovascular disease share aetiologies with dementia, which often has a vascular component. Other comorbidities such as cancers, are independent, co-occurring conditions. Acknowledging this heterogeneity, we investigated how the number of illnesses present at death in people recorded as having dementia varied with area deprivation. Building on previous work by Public Health England [5], we aimed to provide health and social care policymakers and commissioners with data on those dying from dementia. We explored conditions recorded alongside dementia on death certificates and tested the hypothesis that more conditions would be recorded at death in more deprived areas.

## Methods

We analysed death certificates for people aged 18+ at the time of death, obtained from the Office for National Statistics (ONS): Public Health England Annual Mortality Extract. This dataset includes information recorded on the Medical Certificate of Cause of Death (MCCD) from civil registry records, which ONS converts to ICD-10 (International Statistical Classification of Diseases and Related Problems version 10) codes using automatic coding software [6].

We defined the primary (also termed underlying) cause of death as the disease that initiated the train of events directly linked to death; and the contributory cause of death as part of the causal sequence of events contributing to death [5]. Each death certificate includes one primary and up to 15 contributory causes. When a person with a diagnosis of dementia dies from an unrelated cause, dementia may not be listed on the death certificate. Therefore, this report does not describe the whole population who die with dementia; but it enables us to describe those for whom dementia was a cause of death – in other words who died from dementia.

We defined dementia as the presence of any ICD-10 dementia diagnostic code (Table S1). Comorbidity was defined as any condition recorded as a cause of death (primary or contributory) in addition to dementia. We included conditions with ≥5% occurrence in the cohort, categorised using ONS short list of cause of death [6] (Table S2).

We report the number of people in England who died with a dementia diagnosis recorded on their death certificate (as a primary or contributory cause) between 2013 and 2023; the dementia subtype diagnoses, year of death registration; sex; age at death; country of birth and area deprivation, using Index of Multiple Deprivation (IMD) quintiles, with quintile 1 (IMD 1-2) representing the highest socioeconomic deprivation level and quintile 5 (IMD 9-10) the lowest.

We report the proportion of deaths of people with dementia in which each common comorbidity was recorded. We conducted a multiple linear regression analysis to test our hypothesis that number of comorbid disorders was associated with area-level deprivation, after controlling for age and sex.

## Results

Between 2013 and 2023 there were 987 719 deaths recorded with dementia as a cause (primary and contributory), of which 693 663 (70.2%) recorded dementia as the primary cause of death. Consistent with the previous report [5], unspecified dementia (497 416; 50%) was the most commonly recorded subtype, followed by Alzheimer’s disease (256 180; 26%) and vascular dementia (182 527; 19%). The proportion of people dying with unspecified dementia subtype increased with deprivation level, from 47.5% (n=94 760/199 405) of deaths in quintile five to 52.9% (n=95 454/180 523) of deaths in quintile one.

Figure 1 shows the number of deaths with dementia (and subtypes) recorded as a primary or contributory cause of death, by year. The higher number of deaths from dementia in 2020 can be attributed to the Covid-19 pandemic [7].

**Figure 1.**
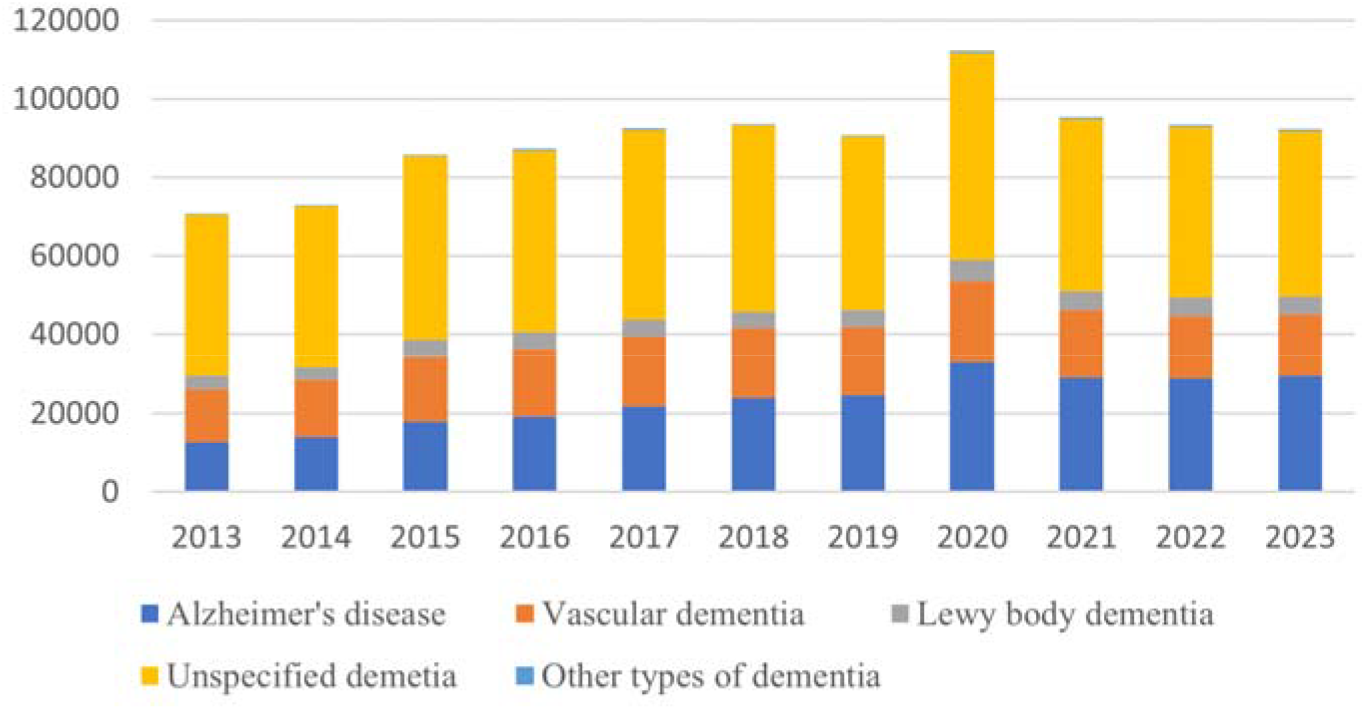
Deaths with dementia as a primary or contributory cause (specifying subtype) by year (N=987, 719)

Of these deaths from dementia (primary or contributory), 614 419 (62.2%) were recorded in women and 373 300 (37.8%) in men. 65% (n= 643 026) were aged 85+. Most were born in England (846 584; 85.7%), Scotland (22 706; 2.3%), Ireland (22 991; 2.3%), Wales (18 166; 1.8%) or India (11 630; 1.1%). These figures are similar to those reported from 2012-14 [5].

### Causes of death

15.1% (n=149 447) of included death certificates recorded dementia as sole cause of death, while others recorded up to 15 comorbidities as primary or contributory causes of death (median=1; IQR=1-2). After controlling for age and sex, death certificates of people living in areas with higher deprivation included more comorbidities (R^2^=0.01, F(6, 987719) =1681, p<0.001) (Table 1).

**Table 1.**
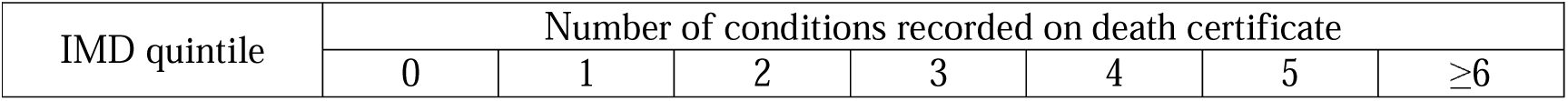
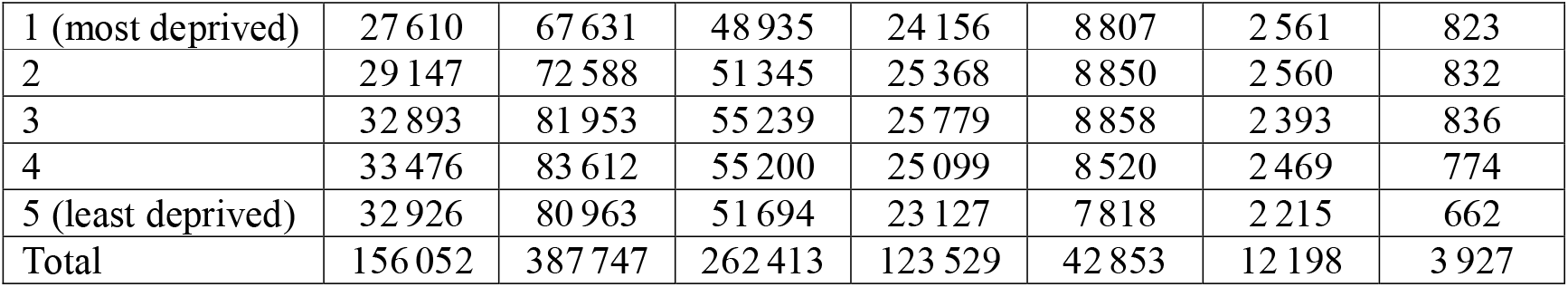
Number of comorbidities recorded on death certificates across deprivation quintiles (1=most deprived)

In cases where dementia was a contributory cause of death, most common primary causes were diseases of the circulatory (107 136; 36.4%), respiratory (31 431; 10.7%) and digestive systems (18 637; 6.3%); and cancers (54 538; 18.5%).

Where dementia was the primary cause, most commonly recorded contributory causes were: ‘senility’ (254 437; 25.7%), influenza and pneumonia (183 203; 18.5%), ischaemic heart diseases (114 871; 11.6%), cancers (107 444; 10.8%), hypertensive diseases (99 517; 10%) and diabetes (98 517; 10%).

## Conclusion

There appears to be a small increase in the number of deaths from dementia from 2013-23, reflecting the aging population [8], pandemic-related excess deaths [7] and increasing dementia diagnosis rates [9]. People living in more deprived areas who died from dementia had more conditions recorded on their death certificates than those who died with dementia in less deprived areas. The associations of socioeconomic deprivation and poor health including premature mortality with higher dementia risk are well-established [10,11,12]. Our results suggest that one reason for this inequity may be the negative sequelae of living with multiple comorbidities. As the number of comorbidities increases, healthcare needs become more complex, especially in people with dementia [13].

In line with previous evidence, we found that people with dementia commonly died with cardiometabolic diseases [14,15], long-term conditions that are implicated in the aetiology of dementia and which affect its prognosis and quality of life [16]. One in ten people dying from dementia had diabetes mellitus. This has implications for end-of-life care, as there are complexities to managing diabetes in people with dementia, including challenges with administering medication, regulating eating habits and regular monitoring of blood glucose [17].

One in ten people who died from dementia had cancer. People diagnosed with dementia at the same time as, or after their cancer diagnosis, are less likely to receive cancer treatment or end-of-life care appropriate to their needs [18]. Our findings highlight the need for improved understanding of the complexities of delivering appropriate care for people with long-term conditions co-existing with dementia [19]. This could help reduce care inequalities.

There are several limitations to the study. Our sample was limited to deaths where dementia was recorded as a significant contributory factor. Death certificates do not provide information on conditions not considered causally related to death. It is less likely dementia will be recorded on death certificates completed in hospital settings. While the accuracy of death certificates as a means of recording dementia diagnosis has increased in recent years, it remains under recorded [20].

Dementia rarely travels alone, especially towards the end of life [21]. Policies to drive higher quality care for people with multiple long-term conditions especially from deprived areas need to ensure that treatment and care, including end of-life care is tailored to complex needs including those posed by dementia as a comorbidity.

## Data Availability

The data is not available

## Acknowledgement

This work is a collaboration between Department of Health and Social Care (DHSC) and Dementia and Neurodegeneration Policy Research Unit-Queen Mary University of London (DeNPRU-QM). This study was conducted as part of a research policy secondment by SZ in the DHSC.

