## Supplementary material for "Association of comorbidities and socioeconomic deprivation among people who died from dementia in England Between 2013-2023: analysis of death certificates"

Supplementary materials

| Table S1: ICD-10 classification codes of the dementia subtypes | |
| --- | --- |
| Condition | **ICD-10 codes** |
| Alzheimer’s disease | F00.0, F00.1, F00.2, F00.9, G30.0, G30.1, G30.8, G30.9 |
| Vascular dementia | F01.0, F01.1, F01.2, F01.3, F01.8, F01.9 |
| Dementia with Lewy bodies and Parkinson’s disease dementia | F02.3, F02.8, G20, G31.8, |
| Unspecified dementia | F03, G31.9 |
| Other types of dementia | F02.0, F02.1, F02.2, F02.4, G31.0, G31.1, G31.2, |

| **Table S2- ONS short list of cause of death** (the chapter on prenatal and pregnancy related disorders was removed) | |
| --- | --- |
| A00 to B99: ICD Chapter I Certain infectious and parasitic diseases | - A00 to A09: Intestinal infectious diseases - A15 to A16: Respiratory tuberculosis - A17 to A19: Other tuberculosis - A39: Meningococcal infection - A40 to A41: Sepsis - B15 to B19: Viral hepatitis - B20 to B24: Human immunodeficiency virus [HIV] disease - B90: Sequelae of tuberculosis |
| C00 to D48: ICD Chapter II Neoplasms | - C00 to C97: Malignant neoplasms - C00 to C14: Malignant neoplasms of lip, oral cavity and pharynx - C15: Malignant neoplasm of oesophagus - C16: Malignant neoplasm of stomach - C18: Malignant neoplasm of colon - C19 to C21: Malignant neoplasm of rectosigmoid junction, rectum and anus - C22: Malignant neoplasm of liver and intrahepatic bile ducts - C23 to C24: Malignant neoplasm of gallbladder and biliary tract - C25: Malignant neoplasm of pancreas - C32: Malignant neoplasm of larynx - C33 to C34: Malignant neoplasm of trachea, bronchus and lung - C43: Malignant melanoma of skin - C44: Other malignant neoplasms of skin - C45: Mesothelioma - C46: Kaposi sarcoma - C50: Malignant neoplasm of breast - C53: Malignant neoplasm of cervix uteri - C54 to C55: Malignant neoplasm of other and unspecified parts of uterus - C56: Malignant neoplasm of ovary - C61: Malignant neoplasm of prostate - C62: Malignant neoplasm of testis - C64: Malignant neoplasm of kidney, except renal pelvis - C67: Malignant neoplasm of bladder - C71: Malignant neoplasm of brain - C81: Hodgkin lymphoma - C82 to C85: Non-Hodgkin lymphoma - C90: Multiple myeloma and malignant plasma cell neoplasms - C91 to C95: Leukaemia - C97: Malignant neoplasms of independent (primary) multiple sites - D00 to D48: In situ and benign neoplasms, and neoplasms of uncertain or unknown behaviour |
| D50 to D89: ICD Chapter III Diseases of the blood and blood-forming organs and certain disorders involving the immune mechanism | - D50 to D64: Anaemias |
| E00 to E90: ICD Chapter IV Endocrine, nutritional and metabolic diseases | - E10 to E14: Diabetes mellitus |
| F00 to F99: ICD Chapter V Mental and behavioural disorders | - F01, F03: Vascular and unspecified dementia - F10 to F19: Mental and behavioural disorders due to psychoactive substance use |
| G00 to G99: ICD Chapter VI Diseases of the nervous system | - G00, G03: Meningitis (excluding meningococcal) - G12.2: Motor neuron disease - G20: Parkinson disease - G30: Alzheimer disease - G35: Multiple sclerosis - G40: Epilepsy |
| H00 to H59: ICD Chapter VII Diseases of the eye and adnexa |  |
| H60 to H95: ICD Chapter VIII Diseases of the ear and mastoid process |  |
| I00 to I99: ICD Chapter IX Diseases of the circulatory system | - I05 to I09: Chronic rheumatic heart diseases - I10 to I15: Hypertensive diseases - I20 to I25: Ischaemic heart diseases - I21 to I22: Acute myocardial infarction - I26 to I51: Other heart diseases - I60 to I69: Cerebrovascular diseases - I60 to I62: Intracranial haemorrhage - I63: Cerebral infarction - I64: Stroke, not specified as haemorrhage or infarction - I70: Atherosclerosis - I71: Aortic aneurysm and dissection |
| J00 to J99: ICD Chapter X Diseases of the respiratory system | - J09: Influenza due to certain identified influenza virus - J10 to J11: Influenza - J12 to J18: Pneumonia - J40 to J44: Bronchitis, emphysema and other chronic obstructive pulmonary disease - J45 to J46: Asthma |
| K00 to K93: ICD Chapter XI Diseases of the digestive system | - K25 to K27: Gastric and duodenal ulcer - K40 to K46: Hernia - K57: Diverticular disease of intestine - K70 to K77: Diseases of the liver |
| L00 to L99: ICD Chapter XII Diseases of the skin and subcutaneous tissue |  |
| M00 to M99: ICD Chapter XIII Diseases of the musculoskeletal system and connective tissue | - M05 to M06, M08: Rheumatoid arthritis and juvenile arthritis - M80 to M81: Osteoporosis |
| N00 to N99: ICD Chapter XIV Diseases of the genitourinary system | - N00 to N15: Glomerular and renal tubulo-interstitial diseases - N17 to N19: Renal failure - N40: Hyperplasia of prostate |
| Q00 to Q99: ICD Chapter XVII Congenital malformations, deformations and chromosomal abnormalities | - Q20 to Q28: Congenital malformations of the circulatory system |
| R00 to R99: ICD Chapter XVIII Symptoms, signs and abnormal clinical and laboratory findings, not elsewhere classified | - R54: Senility - R95: Sudden infant death syndrome - R99: Other ill-defined and unspecified causes of mortality |
| S00 to T98: ICD Chapter XIX Injury, poisoning and certain other consequences of external causes | - S00 to S19: Injuries to the head and the neck - S20 to S29: Injuries to the thorax - S30 to S39: Injuries to the abdomen, lower back, lumbar spine and pelvis - S72: Fracture of femur - T20 to T32: Burns and corrosions - T39.1: Poisoning by 4-Aminophenol derivatives - T40: Poisoning by narcotics and psychodysleptics [hallucinogens] - T42: Poisoning by antiepileptic, sedative-hypnotic and antiparkinsonism drugs - T43: Poisoning by psychotropic drugs, not elsewhere classified - T50.9: Poisoning by other and unspecified drugs, medicaments and biological substances - T51 to T65: Toxic effects of substances chiefly nonmedicinal as to source - T58: Toxic effect of carbon monoxide - T71: Asphyxiation - T75.1: Drowning and nonfatal submersion |
| V01 to Y89 (inc U50.9): ICD Chapter XX External causes of morbidity and mortality | - V01 to X59: Accidents - V01 to V99, Y85: Transport accidents - V01 to V89: Land transport accidents - W00 to W19: Falls - W65 to W74: Accidental drowning and submersion - X00 to X09: Exposure to smoke, fire and flames - X40 to X49: Accidental poisoning by and exposure to noxious substances - X41: Accidental poisoning by and exposure to antiepileptic, sedative-hypnotic, antiparkinsonism and psychotropic drugs, not elsewhere classified - X42: Accidental poisoning by and exposure to narcotics and psychodysleptics [hallucinogens], not elsewhere classified - X44: Accidental poisoning by and exposure to other and unspecified drugs, medicaments and biological substances - X59: Accidental exposure to unspecified factor - X60 to X84: Intentional self-harm [note 1] - X85 to Y09: Assault [note 1] - Y10 to Y34: Event of undetermined intent - X60 to X84, Y10 to Y341: Intentional self-harm; and event of undetermined intent - U50.9, X85 to Y091: Assault; death from injury or poisoning, event awaiting determination of intent (inquest adjourned) |
